# Almond Consumption for 8 Weeks Altered Host and Microbial Metabolism in Comparison to a Control Snack in Young Adults

**DOI:** 10.1101/2021.05.03.21256353

**Authors:** Jaapna Dhillon, John W. Newman, Oliver Fiehn, Rudy M. Ortiz

## Abstract

Almond consumption can improve cardiometabolic (CM) health. However, the mechanisms underlying those benefits are not well characterized. This study explored the effects of consuming a snack of almonds vs. crackers for 8 weeks on changes in metabolomic profiles in young adults (clinicaltrials.gov ID: NCT03084003). Participants (n=73, age: 18-19 years, BMI: 18-41 kg/m2) were randomly assigned to consume either almonds (2 oz/d, n=38) or an isocaloric control snack of graham crackers (325 kcal/d, n=35) daily for 8 weeks. Blood samples were collected at baseline prior to and at 4 and 8 weeks after the intervention. Metabolite abundances in the serum were quantified by hydrophilic interaction chromatography quadrupole (Q) time-of-flight (TOF) mass spectrometry (MS/MS), gas chromatography (GC) TOF MS, CSH-ESI (electrospray) QTOF MS/MS, and targeted analyses for free PUFAs, total fatty acids, oxylipins and endocannabinoids. Linear mixed model analyses with baseline-adjustment were conducted, and those results were used for enrichment and network analyses. Microbial community pathway predictions from 16S rRNA sequencing of fecal samples was done using PICRUST2. Almond consumption enriched unsaturated triglycerides, unsaturated phosphatidylcholines, saturated and unsaturated lysophosphatidylcholines, tricarboxylic acids, and tocopherol clusters (p<0.05). Targeted analyses reveal lower levels of omega-3 total fatty acids (TFAs) overall in the almond group compared to the cracker group (p<0.05). Microbial amino acid biosynthesis, and amino sugar and nucleotide sugar metabolism pathways were also differentially enriched at the end of the intervention (p<0.05). The study demonstrates the differential effects of almonds on host tocopherol, lipid, and TCA cycle metabolism with potential changes in microbial metabolism, which may interact with host metabolism to facilitate the CM benefits.

## INTRODUCTION

The complex interplay among the host genome, the gut microflora, and environmental (diet, lifestyle, socioeconomic determinants etc.) and behavioral factors modulates multiple metabolic pathways resulting in specific biological and physiological effects (1) that ultimately influence health and disease. Metabolomic approaches that integrate large swaths of metabolism provide an experimental advantage by mapping metabolites in such a fashion that putative mechanisms can be elucidated as demonstrated in the SuGAR Project (2–4).

To date, nutritional metabolomics has focused more on the identification of biomarkers for foods and diet-related disease states (5). Over the past decade, potential biomarkers or metabolite signatures have been identified for several foods including red meat (6), coffee (7), citrus fruits (8), cruciferous vegetables (9), almonds (10), and other plant foods (11). However, there is limited research on the elucidation of molecular mechanisms responsible for the effects of dietary interventions (5). The potential of identifying changes in metabolic pathways is improving with advancements in analytical chemistry techniques, metabolite databases, and computational tools.

Another important consideration in nutritional metabolomics is the gut microbiome-host interactions (12). A considerable number of metabolomic features that are influenced by dietary changes can also be modifiable by the gut microbiome. For example, the metabolomic signatures observed in human fecal donor samples were reproduced in the urine and feces of humanized mice (13). Changes to the diets fed to humanized mice altered those metabolomic signatures and mirrored changes in bacterial community composition (13).

Nut consumption is associated with reduced cardiovascular disease mortality (14). Moreover, interventional studies demonstrate favorable effects of nut consumption on clinical lipid profiles, blood glucose, and endothelial function (14). Nut studies employing metabolomics have identified potential urinary biomarkers and signatures of walnut (15) and pistachio (16) as well as urinary (17) and plasma (18) biomarkers of mixed nut consumption and erythrocyte signatures of almond consumption (10). These metabolites collectively include markers of fatty acid metabolism, ellagitannin-derived microbial metabolites, microbial-derived phenolic metabolites, and intermediate tryptophan/serotonin pathway metabolites. There are limited -omics data that map the changes in metabolic pathways following the chronic consumption of nuts. The high unsaturated fat content and relatively low available carbohydrate content of almonds has the potential to change carbohydrate and lipid metabolism.

We have previously demonstrated the effects of almond snacking for 8 weeks on cardiometabolic (19) and microbiome profiles (20) in young adults. The unique nutrient profile of almonds improved postprandial glucoregulation (19) and alpha-diversity of the gut microbiome (20) compared to cracker snacking at the end of the 8-week intervention. Here, we use untargeted and targeted metabolomics to capture the alterations in serum metabolites involved in metabolic pathways with 8 weeks of almond snacking.

## METHODS

All procedures involving human subjects were approved by the University of California (UC) Merced Institutional Review Board. The study is registered on ClinicalTrials.gov (registration number: NCT03084003). The samples used in the present study were collected previously and presented separately here to provide the requisite focus on these results, which complement our previous studies (21, 22)

### Participants

Seventy-three (41 women and 32 men) young adults (18–19 years old, BMI: 18-41 kg/m^2^) were recruited to participate in an 8-week randomized, controlled, parallel-arm intervention examining the effects of almond vs. cracker snacking on cardiometabolic, microbiome, and metabolomics outcomes. The eligibility criteria were as follows: (a) 18-19 years of age, (b) newly enrolled, 1st-year college students with no nut allergies, (c) non-smokers, and (d) no diagnosed cardiometabolic disorders. Participants were recruited via advertisements and those who met eligibility criteria provided written, informed consent before beginning the study.

### Study design and protocol

The primary study design has been described previously (19). The sample size calculations for the primary study were based on glucoregulatory profiles at the end of the 8-week intervention (19). Participants were randomized into one of two study arms: (1) almonds and (2) crackers. Participants in the almond group (n=38) consumed 57 g/d (2 oz; 327 kcal; 14% carbohydrate (8g fiber), 74% fat, 13% protein) of whole, dry-roasted almonds. Participants in the cracker group (n=35) served as the isocaloric control consuming 5 sheets (77.5 g/d) of graham crackers (325 kcal; 74% carbohydrate (2.5 g fiber, 20% fat, 6% protein). The cracker group was asked to avoid all nuts, seeds, and nut-containing products. Anthropometric, biochemical, dietary and microbiome data were collected and analyzed as described previously (23). Serum samples were collected at baseline prior to starting the intervention and at 4 and 8 weeks as previously described (21) and stored at −80 °C.

### Gas chromatography (GC) time-of-flight (TOF) mass spectrometry (MS) data acquisition and processing

Serum aliquots were thawed, extracted, trimethylsilyl-derivatized, and the metabolite abundances quantified by GCTOF MS as previously described (24). Derivatized samples were analyzed on an Agilent 7890 A gas chromatograph (Santa Clara, CA) equipped with a 30 m x 0.25 mm i.d., 25 μm Rtx5Sil-MS column with an additional 10 m integrated guard column (Restek, Bellefonte PA) (25, 26). Mass spectrometry was performed using a Leco Pegasus IV time-of-flight mass spectrometer (St. Joseph, MI). Thirteen internal standards, C8–C30 fatty acid methyl esters, were added to samples for retention time alignment markers, quality control purposes and quantification corrections.

Raw mass spectra were preprocessed using ChromaTOF vs. 4.0 and were further processed using the BinBase algorithm and database for metabolite identification (27, 28). Quantification of metabolites are reported as peak heights.

All samples were analyzed in one batch, and data quality and instrument performance were constantly monitored using quality controls comprised of pooled serum samples and injected every 10 samples.

### Hydrophilic Interaction chromatography (HILIC) quadrupole (Q) time-of-flight (TOF) MS/MS data acquisition and processing

Serum aliquots were extracted as previously described (29). The bottom layer of the 2-phase solution was used for HILIC-MS. Extracts were analyzed on a Agilent 1290 Infinity II LC System (Santa Clara, CA) with a 150 mm long, 2.1 mm interdiameter (id), and 1.7 μm particles Waters Acquity UPLC BEH Amide column protected by a short guard column. Mass spectrometry was performed using a Sciex 6600 TTOF mass spectrometer (Framingham, MA) with resolution R=10,000 for positively charged polar compounds. Nineteen internal standards, optimized for HILIC-MS, were added to samples for retention time alignment markers, quality control purposes, and quantification corrections.

Raw data were processed using MS-DIAL (version 3.2) (30). Quantification of metabolites are reported as peak heights. All samples were analyzed in one batch, and data quality and instrument performance were constantly monitored using blanks and quality controls, which were comprised of pooled plasma samples (BioIVT) and injected every 10 samples.

### Charged Surface Hybrid electrospray (CSH-ESI) QTOF MS/MS data acquisition and processing

Serum aliquots were extracted as previously described (29). The top layer of the 2-phase solution was used for lipidomic analyses. Extracts were analyzed on an Agilent 1290 Infinity II LC System (Santa Clara, CA) with a 100 mm long, 2.1 mm id, and 1.7 μm particles Waters Acquity UPLC CSH C18 column protected by a short guard column. Mass spectrometry was performed using an Agilent 6530 QTOF mass spectrometer with resolution R=10,000 for positively charged lipids and an Agilent 6530b QTOF mass spectrometer with resolution R=20,000 for negatively charged lipids. Twenty-four internal standards, optimized for lipidomics, were added to samples for retention time alignment markers, quality control purposes, and quantification corrections.

Raw data were processed using MS-DIAL (version 2.8) (30). Quantification of metabolites are reported as peak heights. All samples were analyzed in one batch, and data quality and instrument performance were constantly monitored using quality controls, which were comprised of pooled plasma samples (BioIVT) and injected every 10 samples.

### Targeted analyses of total fatty acids

Serum aliquots were extracted, derivatized, and analyzed as previously described (31, 32). Extracts in 8:1 methanol/toluene were transformed into fatty acid methyl esters (FAMEs) by sequential incubation with methanolic sodium hydroxide and methanolic hydrochloric acid, isolated in hexane from neutralized solutions and quantified by GC-MS. Briefly, FAMEs were separated on a 30m x 0.25mm, 0.25 µm DB-225ms on a 6890 gas chromatogram interfaced with a 5973A mass selective detector (Agilent Technologies) and quantified against 6-to 8-point calibration curves. Data was acquired with Chemstation v E.02 and processed with MassHunter v. 3.0.2. Results were corrected for recoveries of perdeuterated palmitate introduced as a triglyceride prior to extraction.

### Targeted analyses of free PUFAs, oxylipins, and endocannabinoids

Serum aliquots were extracted as previously described (33). Residues in isopropanol extracts were separated on a 2.1 mm x 150 mm, 1.7 µm BEH C18 column (Waters, Milford, MA) and detected by electrospray ionization with multi reaction monitoring on a API 6500 QTRAP (Sciex; Redwood City, CA). Metabolites were quantified against 7-to 9-point calibration curves of authentic standards and internal standard corrections using modifications as previously reported (2).

### Univariate analyses of metabolomics data

Data were reported as quantitative ion peak heights and known metabolites were normalized using Systematic Error Removal Using Random Forest (SERRF) normalization (34). SERRF consistently produced the lowest cross-validated relative standard deviation (cvRSD) for QC samples compared to 14 other commonly used normalization methods (34). Metabolites with QC RSD > 50% were excluded from further analyses. For HILIC-MS, metabolite intensities in samples were also compared to blanks. Metabolites that showed no significant differences between sample and blank (i.e. Wilcoxon test p-value > 0.05) and those with a median sample to blank ratio < 1 were also excluded from further analyses. Data not meeting normality assumptions were transformed using Johnson’s family of transformations. However, only non-transformed data and means are presented for interpretation of biological significance.

We took a 2-step approach to the analyses. The first step focused on selecting the identified metabolites that showed a significant overall time effect in a linear, mixed model analysis with week (baseline, week 4, and week 8) as the factor. The time effect p-values for metabolites were corrected for multiple hypotheses testing using Benjamini-Hochberg correction (false discovery rate (FDR) adjusted p-value). Metabolites that demonstrated a significant (FDR < 0.05) time effect were selected for further analyses. The second step comprised of the selected metabolites undergoing a linear, mixed model analysis with week (week 4 and week 8), snack group, and a week-by-snack group interaction as factors where the analyses were adjusted for baseline values. When significant interaction effects were observed, simple effects for time and snack were carried out and pairwise comparisons were adjusted for multiple comparisons using Bonferroni correction. For any correlation analyses by groups, the significance of the difference between correlation coefficients was assessed using Fisher r to z transformation. Future analyses will explore the effects of BMI, sex, and metabolic risk factors which are outside the scope of this manuscript.

### Chemical enrichment analysis

Chemical similarity enrichment analysis was conducted using ChemRICH, which is a software for metabolomic datasets that uses medical subject headings and Tanimoto substructure chemical similarity coefficients to cluster metabolites into non-overlapping chemical groups (35). The quantitative data set comprised of the baseline-adjusted overall snack effect p-values of all annotated metabolites. Statistically significant p-values for clusters of metabolites were obtained by self-contained Kolmogorov–Smirnov tests and adjusted for FDR.

### Network analysis

Network analysis was used to explore differences between the almond and cracker groups within a biochemical and structural context. A biochemical and chemical similarity network was created for all measured metabolites with KEGG and PubChem CID identifiers using MetaMapR (36). Metabolites involved in biochemical transformations were connected based on product-precursor relationships defined in the KEGG RPAIR database. Metabolites sharing structural properties defined in PubChem Substructure Fingerprints (37) were connected at a Tanimoto similarity threshold ≥ 0.7. The quantitative data set comprised of the overall effect size (Hedge’s g for almond vs cracker groups) and baseline-adjusted snack effect p-values. In cases of significant time x group effects, the larger magnitude of effect size between week 4 or week 8 was presented. Since this was an exploratory analysis, the p-values were not FDR adjusted. The network was then visualized in Cytoscape 3.7.2 (38) using the yFiles organic layout and visual separation of clusters in the network was facilitated with the GLay community clustering algorithm (39). Significant metabolites which did not have KEGG identifiers are included as independent nodes with manually annotated edges in their respective pathway clusters.

### Microbial community metabolic potential

The functional potential of the microbial community was predicted from 16S rRNA using Phylogenetic Investigation of Communities by Reconstruction of Unobserved States (PICRUSt2) (40, 41), which uses hidden-state prediction algorithms to infer gene families (i.e. KEGG orthologs (KO) here) and MinPath for inference of pathway abundances. The gene abundances were analyzed by constructing a combined time x group factor and fitting the within-subject correlation in a linear model blocked on subject in the limma-voom pipeline in R (42, 43). Pre-planned contrasts were constructed from the model. KO enrichment of differentially expressed (p<0.05 corrected for multiple contrast testing) metabolic pathway genes for each contrast were conducted in MicrobiomeAnalyst (44) via hypergeometric tests to evaluate if specific metabolic pathways are represented more often than expected by chance. P-values for pathway enrichment were adjusted for FDR.

We identified the serum metabolites potentially derived from gut microbial metabolism using the Annotation of Metabolite Origins via Networks (AMON) tool (45). Participants gut microbiomes were profiled previously via 16S rRNA sequencing of stool samples (46). The input dataset submitted to AMON comprised of microbiome KO identifiers, KO list of the human genome, and KEGG compound IDs of the metabolomics data. The metabolites produced or metabolized by gut microbiome are annotated in the networks (**Figure 1**). The contribution of the microbial community metabolism to the serum metabolite abundances was evaluated using MIMOSA2 (47, 48).

**Figure 1:**
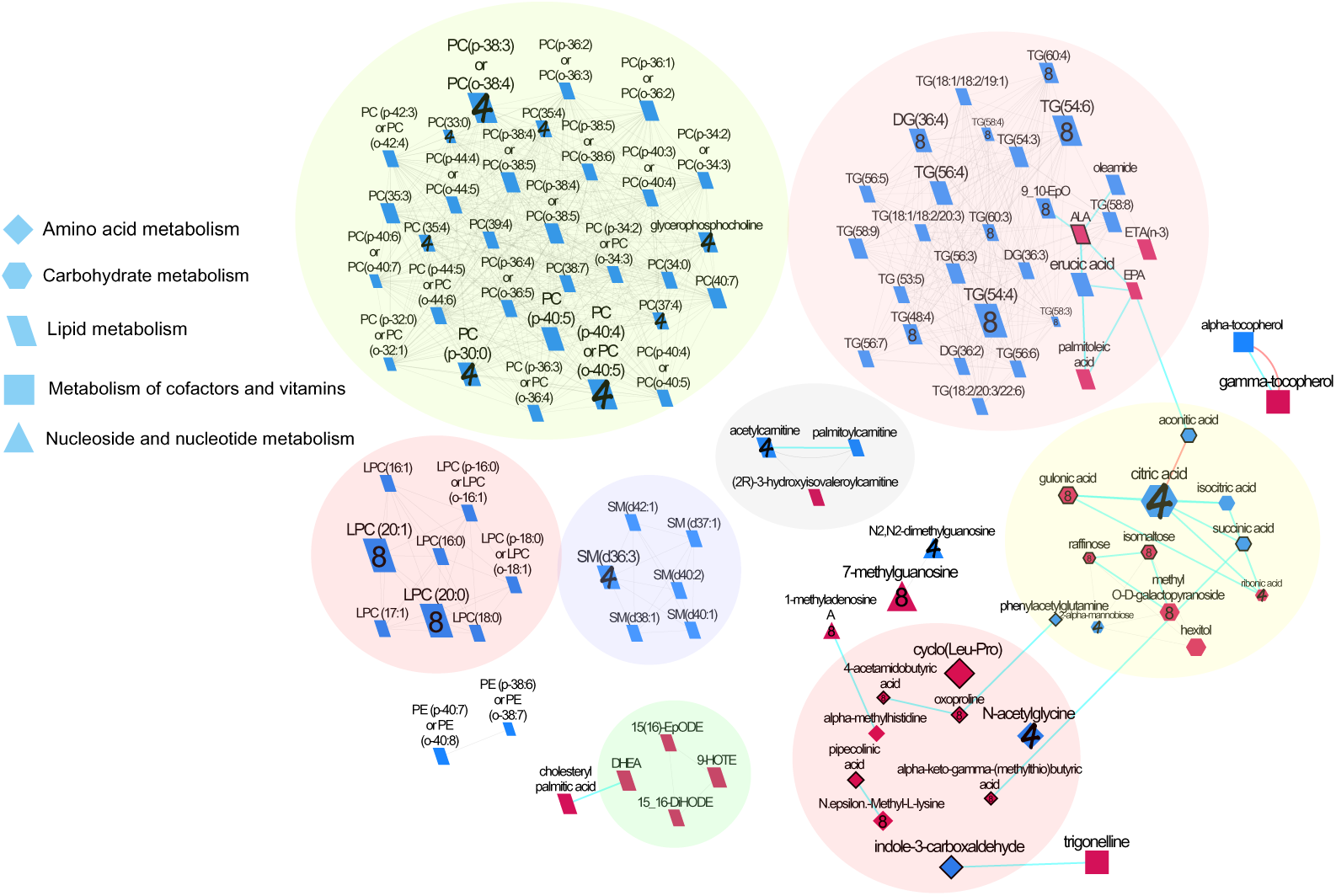
Biochemical network displaying differences between almond and cracker groups over 4 and 8 weeks. Metabolites are connected based on biochemical relationships (orange, KEGG RPAIRS), measured structural similarity (blue, Tanimoto coefficient ≥ 0.7), or manually annotated structural similarity (grey). Metabolite size denotes the effect size (Hedge’s g, almond vs cracker group). Metabolite color represents the direction of the effect size i.e. blue, almond > cracker overall (P<0.05); red, almond < cracker overall (P<0.05). Numbers denote the time point at which the significant i.e. (P<0.05) time x snack effect was observed i.e., 4, week 4; 8, week 8; no number represents significant overall snack effect. P-values are derived from the baseline-adjusted linear mixed model analysis. Shapes display primary metabolic pathways or structural superclass designations obtained via ClassyFire. Metabolites potentially produced by the gut microbiome are highlighted with thick black borders. Clusters of metabolites are circled. Significant metabolites which did not have KEGG identifiers are included as independent nodes with manually annotated edges in their respective pathway clusters.

## RESULTS

### Participant characteristics and findings from parent study

Participant demographic, clinical and dietary characteristics (19) and gut microbiome profiles (46) have been described in detail previously.

### SERRF normalization reduced the cvRSD of QCs in the untargeted analyses

For GC-TOF MS, the cvRSD of QCs decreased from 23.7% (raw) to 12.7% (SERRF). For ESI (-) QTOF-MS, cvRSD of QCs decreased from 14% (raw) to 6.5% (SERRF). For ESI (+) QTOF-MS, cvRSD of QCs decreased from 17.9% (raw) to 4.5% (SERRF). For HILIC-MS, cvRSD of QCs decreased from 15% (raw) to 10% (SERRF).

### Data quality of the targeted total fatty acid, oxylipin, endocannabinoid, and non-esterified PUFA analyses

Surrogate recoveries were between 28 - 108% for all oxylipin and endocannabinoid analytes and 54% for fatty acids. Analytical precision was assessed by duplicate analysis of a plasma pool control (UTAK) (n=20) and was excellent with 83% of oxylipin and endocannabinoid analytes with <30% RSD and 63% of fatty acids with <30% RSD for the UTAKs analyzed, respectively.

### Metabolomic coverage

A total of 542 metabolites were quantified by GC-MS of which 150 had known PubChem identifiers, 2747 were quantified by QTOF-MS of which 470 had known PubChem identifiers, and 2427 were quantified by HILIC-MS of which 232 had known PubChem identifiers. The targeted analyses detected and quantified 23 total fatty acids, 29 oxylipins, 10 endocannabinoids, and 5 non-esterified PUFAs above the limits of detection. In total, 692 metabolites were uniquely identified after removal of duplicates and quality checks.

### Top metabolites

A total of 129 untargeted metabolites had significant (FDR<0.05) overall time effect. The means of those untargeted metabolites that had baseline-adjusted snack and snack x time p-values <0.05 are shown in **Table 1**. Almond consumption resulted in overall greater levels of alpha-tocopherol, ecgonine, erucic acid, indole-3-carboxaldehyde, lysophosphatidylcholines (18:0), and oleamide, and lower levels of 3-hydroxyisovaleroylcarnitine and CE (16:0) than cracker consumption (baseline-adjusted snack effect, p<0.05).

**Table 1:**
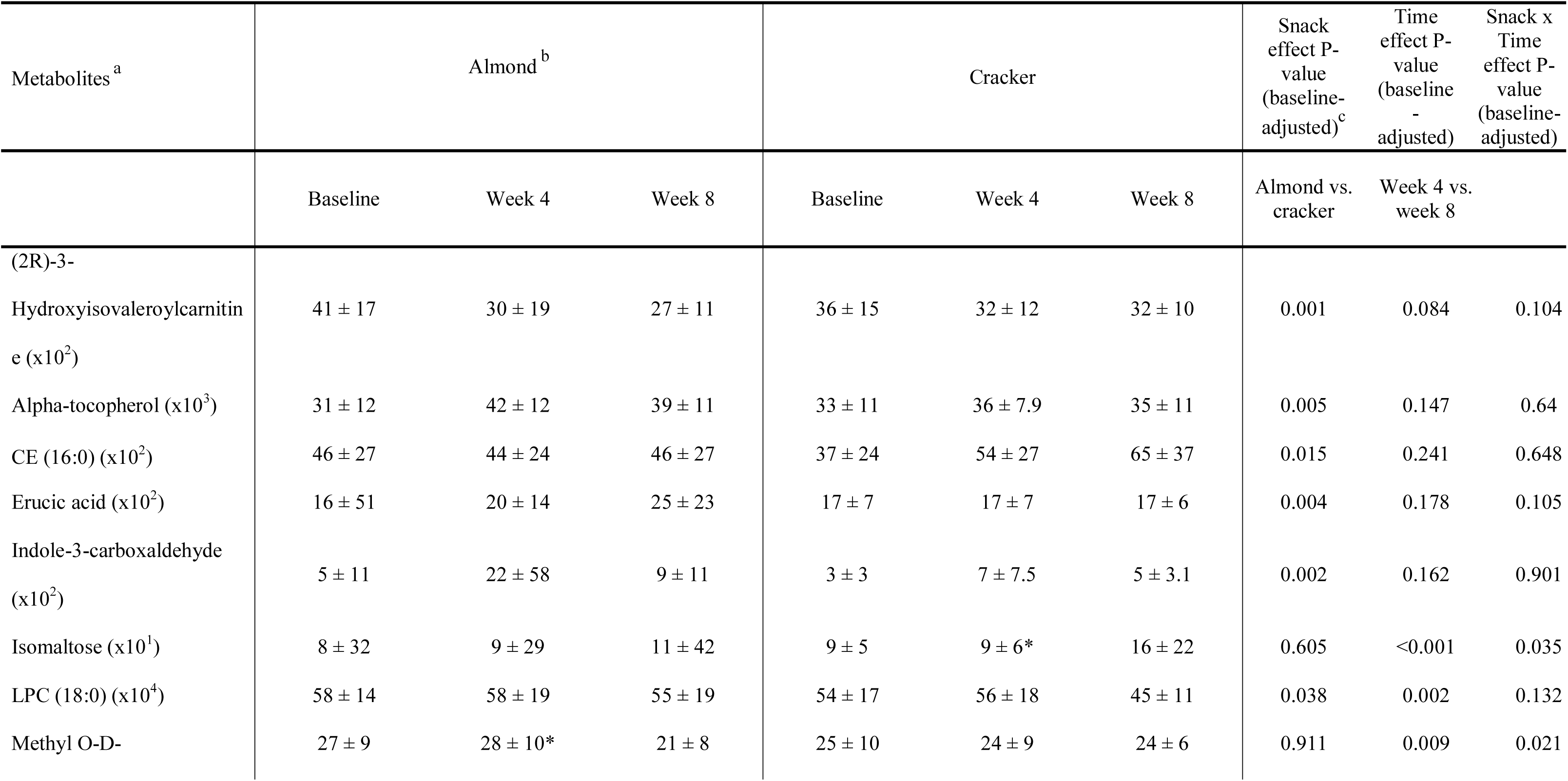
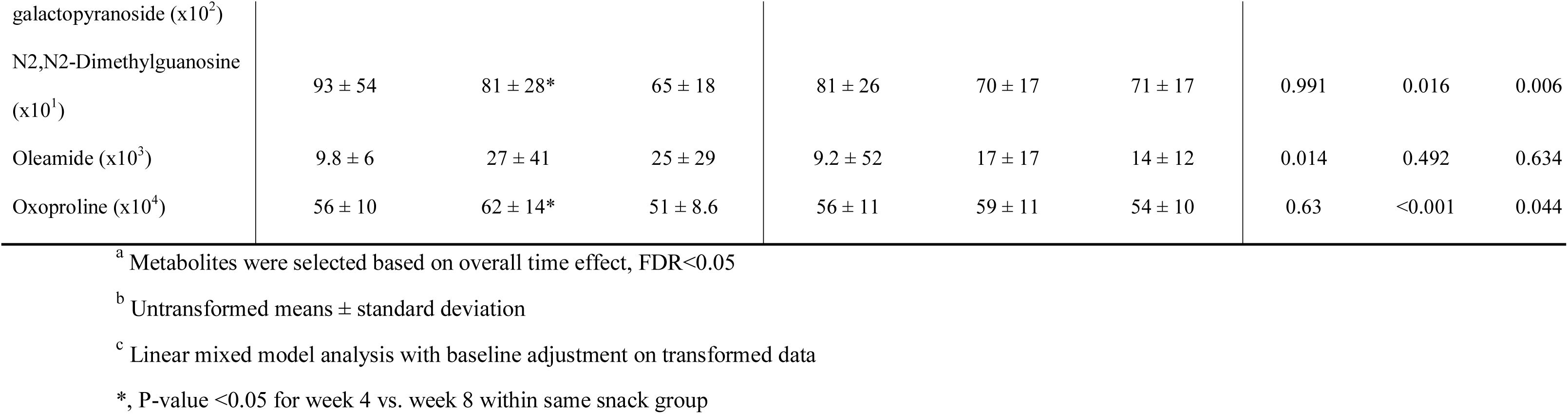
Baseline-adjusted least square means of selected metabolites following almond and cracker consumption over 8 weeks

Several metabolites depicted significant snack and time interactions (baseline-adjusted snack x time effect, p<0.05, **Table 1**). Contrasts depict that cracker consumption resulted in greater levels of isomaltose at week 4 compared to week 8 (p<0.05). Almond consumption resulted in greater levels of oxoproline, methyl O-D-galactopyranoside, and N2,N2-Dimethylguanosine at week 4 compared to week 8 (p<0.05).

The means of specific targeted metabolites that showed significant (p<0.05) baseline-adjusted snack and snack x time effects are shown in **Table 2**. Almond consumption resulted in overall lower levels of NEFA and TFA fractions of alpha-linolenic acid (ALA) and eicosapentaenoic acid (EPA), and eicosatetraenoic acid (ETA) n3 NEFA, and ALA metabolites, 9-HOTE, 15(16)-EpODE, and docosahexaenoic acid (DHEA) compared to cracker consumption (baseline-adjusted snack effect, p<0.05). The omega-6:omega-3 and DPA:EPA fatty acid ratios were higher, and Σ omega-3 and palmitoleic acid:palmitic acid ratio were lower in response to almond consumption compared to cracker consumption (baseline-adjusted snack effect, p<0.05).

**Table 2:**
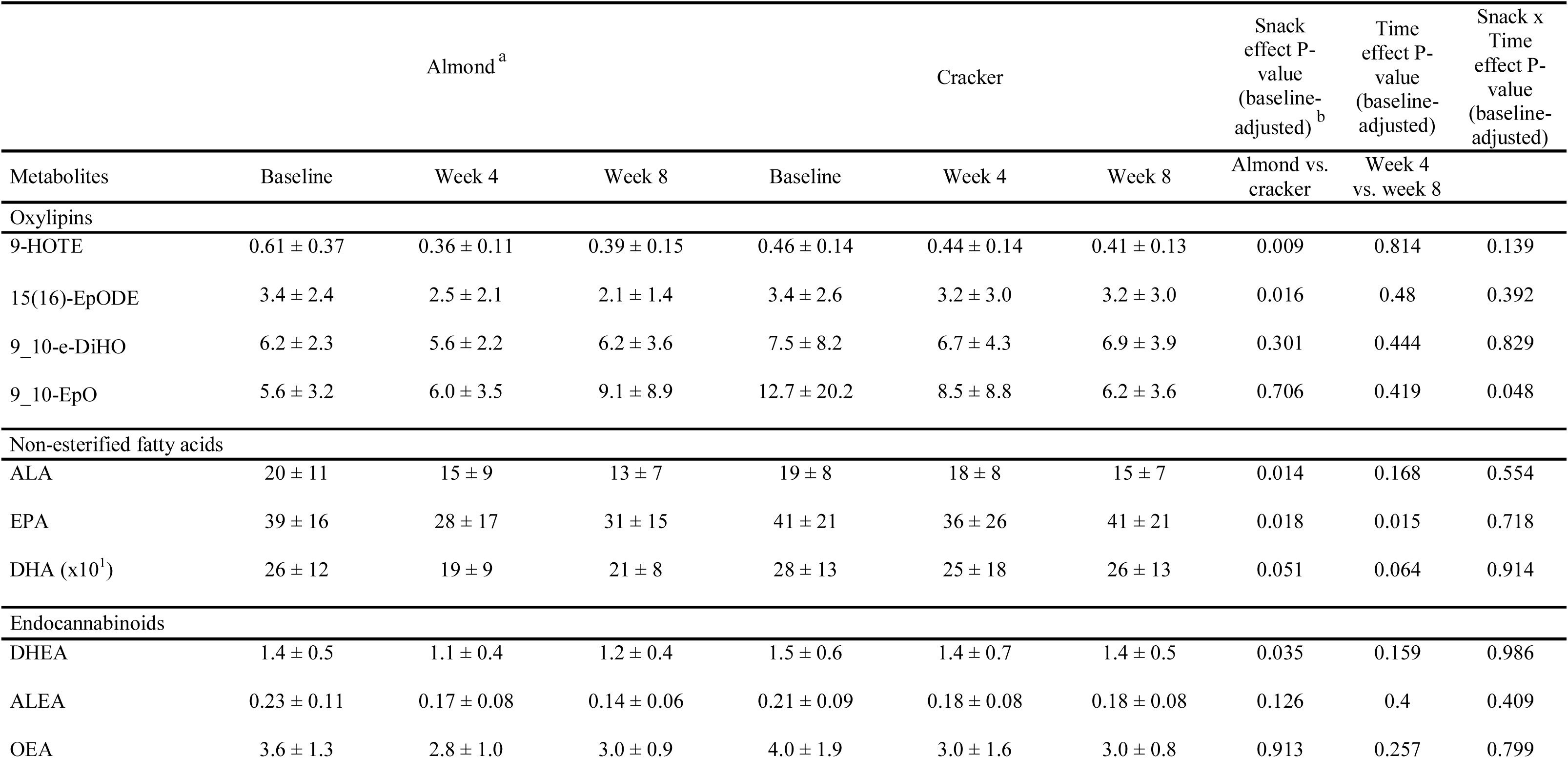
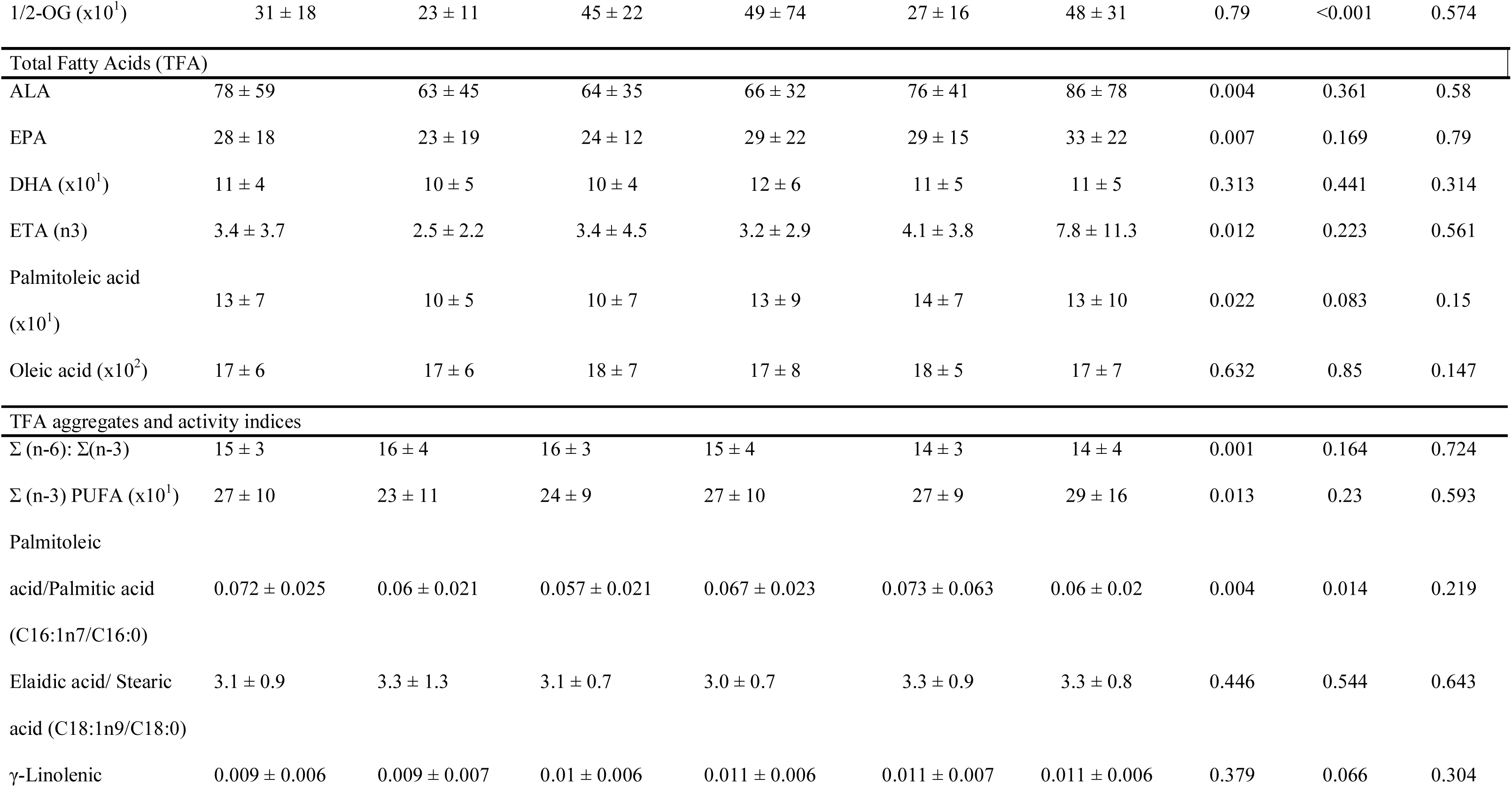
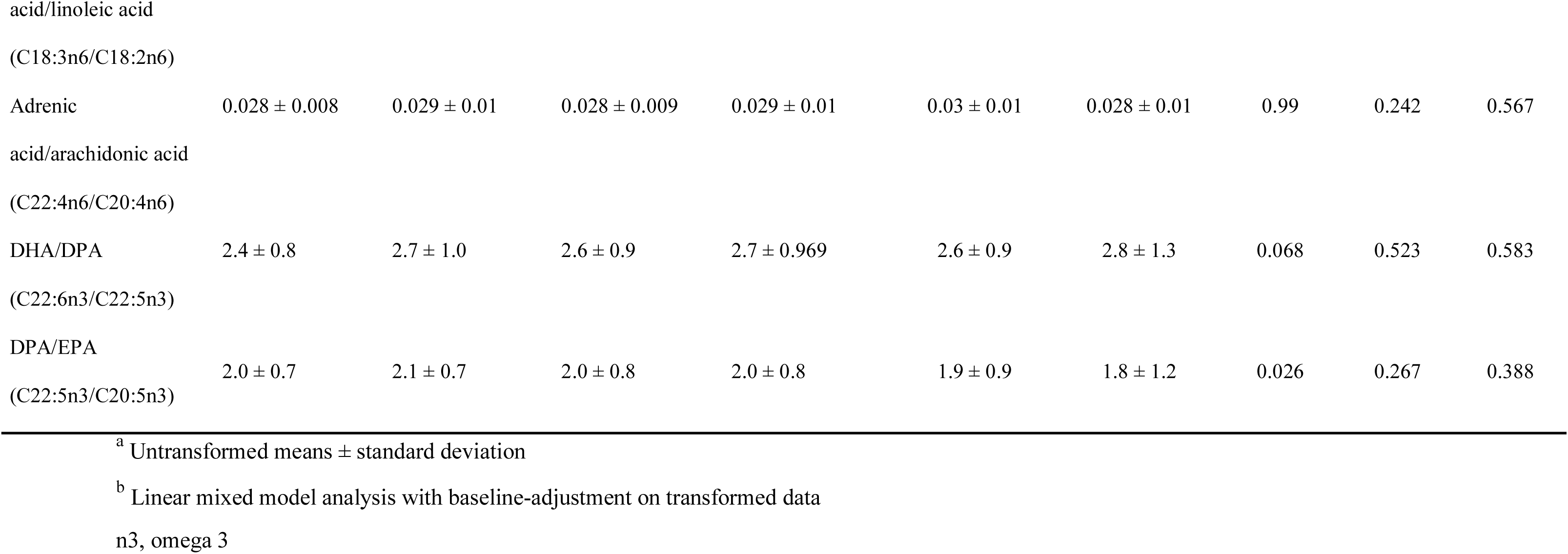
Specific targeted metabolites and their ratios following almond and cracker consumption for 8 weeks

### Chemical similarity enrichment analysis of metabolites indicates greater changes in unsaturated triglycerides and lysophosphatidylcholine metabolism with almond consumption

ChemRICH analysis, which was conducted on baseline-adjusted metabolite values, mapped 625 of the identified (untargeted) metabolites to 58 non-overlapping chemical classes. Seven clusters were significantly different between the almond and cracker groups (FDR-adjusted p-value < 0.05) overall. Almond consumption enriched unsaturated triglycerides, unsaturated phosphatidylcholines, saturated and unsaturated lysophosphatidylcholines, sphingomyelins, and tricarboxylic acids clusters. The key metabolites in each of those clusters are depicted in **Table 3**. The tocopherol cluster was also influenced with almond consumption overall (FDR-adjusted p-value <0.05) with gamma-tocopherol significantly decreased (p<0.05) and alpha-tocopherol increased compared with cracker consumption (**Table 3**).

**Table 3:**
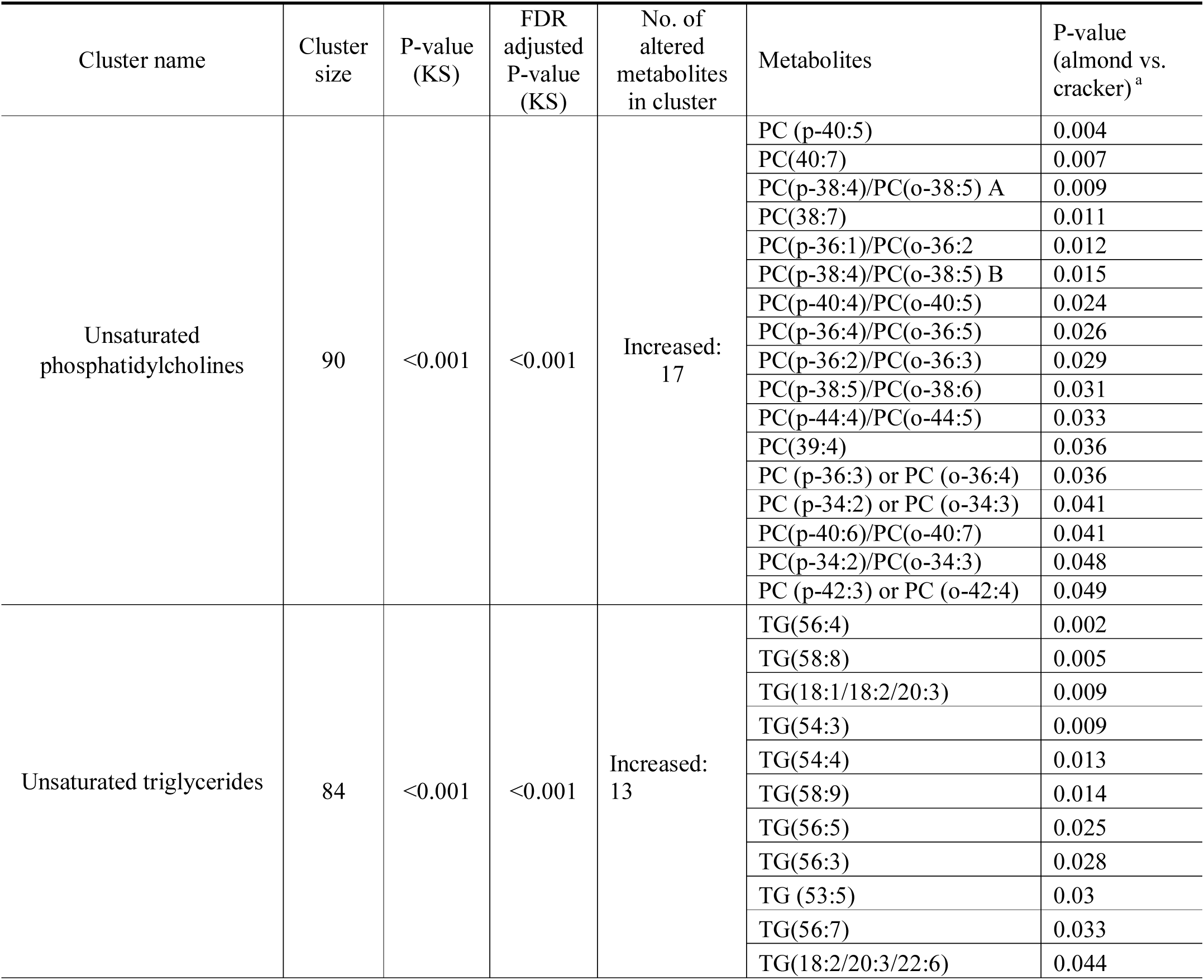
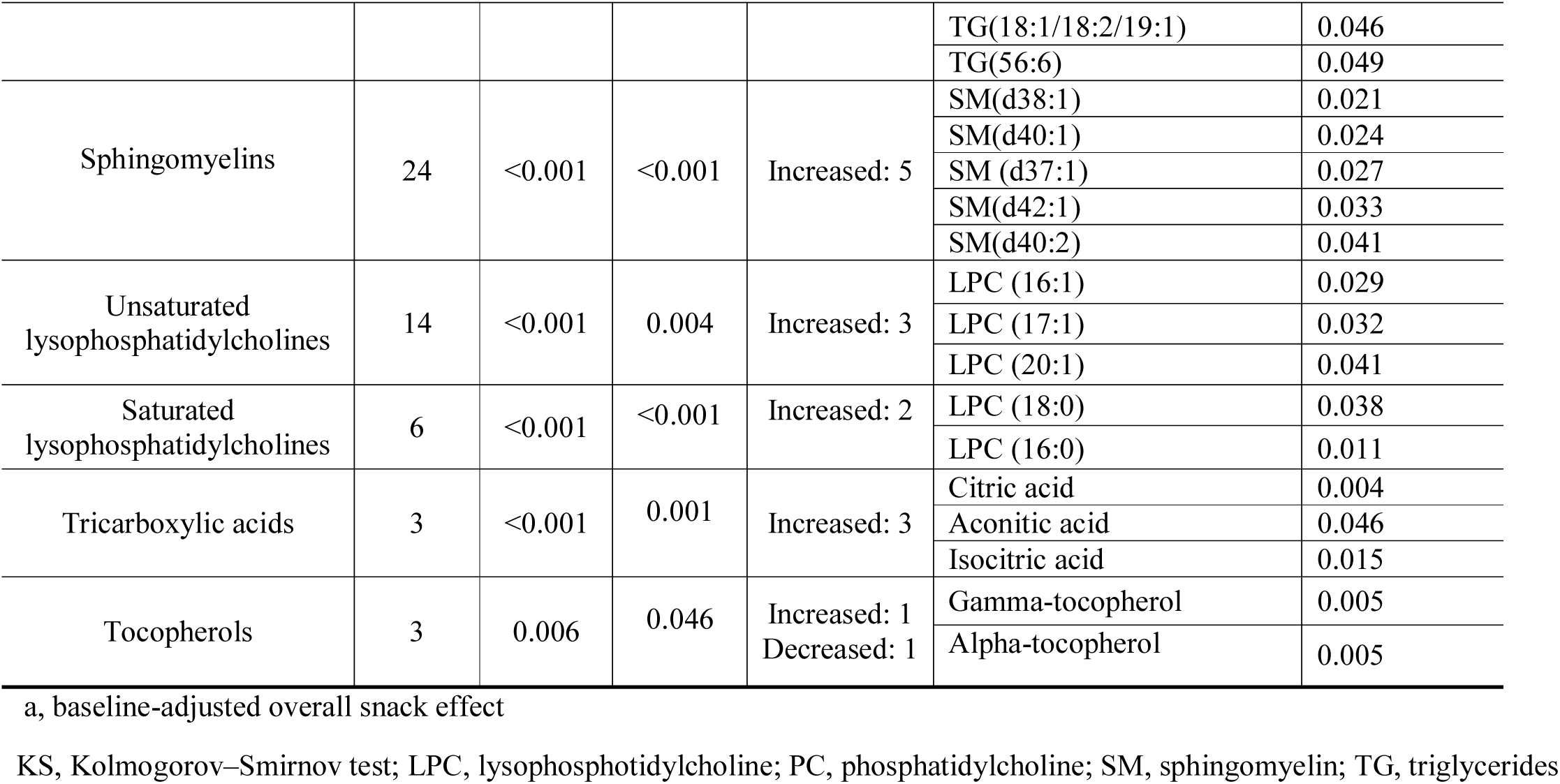
Chemical similarity enrichment analysis results of significant clusters of untargeted serum metabolites measured following almond and cracker consumption over 8 weeks

### Network analysis of metabolites depicts an alteration in compounds involved in lipid and tocopherol metabolism in response to almond consumption

Metabolites were clustered into groups based on biochemical and structural similarities. Network clusters were largely comprised of primary and intermediate metabolites involved in amino acid, carbohydrate, lipid, and nucleotide metabolism (**Figure 1**). More specifically, clusters of amines/amides, TCA cycle metabolites, and carnitine and tocopherol metabolism were also identified (**Figure 1**).

### Carbohydrate metabolism

The almond group had greater levels of metabolites involved in the TCA cycle such as succinic, aconitic, and isocitric acids, and lower levels of hexitol compared to cracker consumption (Hedge’s g: 0.34-0.59, snack effect p<0.05). Almond consumption resulted in greater levels of citric acid at week 4 (g=0.84) and lower levels of raffinose (g=0.26), isomaltose (g=0.32), gluconic acid (g=0.48), methyl O-D-galactopyranoside (g=0.41), and 2-alpha-mannobiose (g=0.28) at week 8 and lower levels of ribonic acid at week 4 (g=0.24) compared to cracker consumption (time x snack effect, p<0.05).

### Amino acid metabolism

Almond consumption largely elicited lower levels of metabolites involved in amino acid metabolism such as cyclo (Leu-Pro), alpha-methyl-histidine, and pipecolinic acid (g=0.31-0.65) and higher levels of phenylacetylglutamine (g=0.27) and indole-3-carboxaldehyde (g=0.56) compared to cracker consumption (snack effect, p<0.05). Differential time x snack effects (p<0.05) were noted as well with 4-acetamidobutyric acid, alpha-keto-gamma-(methylthio) butyric acid, and oxoproline lower with almond consumption at week 8 (g=0.27-0.39) but not at week 4. N-acetylglycine was greater in the almond group at week 4 (g=0.61) and N.epsilon.-methyl-L-lysine was lower in the almond group at week 8 (g=0.41).

### Lipid metabolism

The alterations in lipid metabolites such as LPC, PC, TG, and SM, and fatty acids and their derivatives with almond consumption have been described in the previous sections. Other significant metabolites include 3-hydroxyisovaleroylcarnitine (g=0.36) and palmitoylcarnitine (g=0.31), which was increased in the almond group (snack effect, p<0.05). Differential time x snack effects (p<0.05) were noted for acetylcarnitine (g=0.47) and glycerophosphocholine (g=0.12), both being greater with almond consumption at week 4. LPC (20:0) and LPC (20:1) were greater in the almond group at week 8 (g=0.72-0.76, time x snack effect p<0.05).

### Metabolism of cofactors and vitamins

Almond consumption elicited greater levels of alpha-tocopherol (g=0.49) and lower levels of gamma-tocopherol (g=0.53) and trigonelline (g=0.56) overall compared to cracker consumption (snack effect p<0.05).

### Nucleoside and nucleotide metabolism

Almond consumption elicited lower levels of 7-methylguanosine overall (g=0.37, snack effect p<0.05). Differential time x snack effects (p<0.05) were noted for 1-methyladenosine A and N2,N2-dimethylguanosine with greater levels at week 4 (g=0.29-0.45) and lower levels at week 8 for the almond group (g = 0.32).

### Potential microbiome community-dependent metabolism

There were no significantly enriched pathways detected for the differentially expressed (almond vs. cracker) genes at baseline (FDR>0.05, **Supplemental Table 1**). Overrepresentation analysis of the differentially expressed metabolic pathway genes at week 8 found significant enrichment (FDR<0.05, **Supplemental Table 1**) of biosynthesis of amino acids, amino sugar and nucleotide sugar metabolism, starch and sucrose metabolism, and fructose and mannose metabolism.

AMON identified 94 metabolites in the serum as being potentially produced or metabolized by microbes isolated from 16S sequencing of host stool samples, which are annotated in **Figure 1**. MIMOSA predicted the microbial community potential (CMP) of 81 metabolites out of which 67 overlapped with AMON results. The almond group at week 8 demonstrated a positive correlation of serum N-acetyl-D-mannosamine (R-square: 0.14, p<0.05) (**Supplemental Figure 1**), and a negative correlation of serum L-cysteine (R-square: 0.16, p<0.05) with the CMP scores, which were significantly different from the cracker group (p<0.05 for difference in correlation coefficients between groups). The cracker group at week 8 only demonstrated a negative correlation of 5(S)-HETE (R-square: 0.15, p<0.05) with the CMP scores. The negative association between a metabolite’s levels and its CMP could potentially be due to missing reactions in the KEGG database or effects other than metabolism (48).

## DISCUSSION

The metabolomics analyses in our study demonstrated shifts in host metabolism with almond consumption particularly that of tocopherol, lipids, and the TCA cycle with some differential time effects noted as well. The study also suggests that changes in microbial metabolism could potentially influence host metabolism.

Almond consumption for 8 weeks increased alpha-tocopherol and decreased gamma-tocopherol, which is supported by targeted studies (49). This could be due to the preferential uptake of alpha-tocopherol in the liver when dietary intake is increased and reduced retention of other tocopherol forms (50). Increased alpha-tocopherol is typically considered a biomarker for compliance with almond consumption. The increase in alpha-tocopherol was most prominent at week 4 and appeared to have plateaued at week 8 suggesting that levels may reach a saturable limit by week 4. Previous studies have demonstrated that in participants with normal plasma alpha-tocopherol concentrations of 25 µM, the plasma concentrations do not increase more than 2-3 fold even upon supplementation (50–52) further suggesting that levels of alpha-tocopherol are saturable. In a dose-dependent almond study, 10% of energy intake from almonds resulted in a 13.7 % increase, and 20% in 18.7 % increase in plasma alpha-tocopherol over 4 weeks (49). In our study, the decrease in the magnitude of alpha-tocopherol concentration change (over 8 weeks) as baseline concentrations increase also suggests the presence of a saturable pool (**Supplemental Figure 2**). Our dietary recall data also indicates an increase in alpha-tocopherol over 4 weeks with no further change beyond week 4 with almond consumption (21). Given the differential response in alpha- and gamma-tocopherol to almond consumption, we propose that the ratio may serve as a robust marker of dietary compliance when the intervention may be of a lesser quantity and/or duration to not reach saturable levels in circulation.

Almond consumption was also associated with greater levels of phosphatidylcholines, and its derivative LPC species, which have been reported to influence diverse cell types such as endothelial cells, adipocytes, hepatocytes, and immune cells (53). Due to the complexity of its metabolism, the role of LPCs in disease causality is controversial (53). In a recent review (53), while *in vitro* studies implicated LPCs in apoptosis and pro-inflammatory conditions, clinical studies demonstrated that circulating LPCs were inversely associated with cardiovascular diseases. Other studies showed lower levels of diverse LPC species with obesity (54) and negative associations with inflammatory markers and insulin resistance (54, 55) suggesting that LPCs may protect against metabolic disorders. Analyses conducted on a subset of participants in the current study demonstrated significant (p<0.05) negative correlations of LPCs with inflammatory markers. Moreover, almond consumption for 8 weeks elicited positive correlations (r=0.30-0.55) of IL-6 with various LPC species including LPC (20:1), LPC (18:0), and LPC (18:1), and IL-10 with LPC (22:4) whereas negative correlations (r=0.2-0.41) were identified in the cracker group (p<0.05 for group comparisons). IL-10 is an anti-inflammatory cytokine while IL-6 can be either inflammatory or anti-inflammatory (56). These relationships suggest that LPCs and inflammatory markers in relatively healthy adults are mediated by dietary conditions and that chronic almond consumption may have a role to play in immune health through the maintenance or increase in LPCs. However, other considerations for interpretation of LPC effects across diet studies should include examination of tissues and/or cell types, and comparisons between healthy and diseased states and/or saturated and unsaturated species.

Almond consumption generally increased unsaturated triglycerides over 8 weeks. More specifically, the almond group had higher levels of oleamide, which is a fatty acid amide of oleic acid. Surprisingly, the targeted analyses did not detect a difference in oleic acid TFA between groups even though it is the most predominant fatty acid in almonds. However, this inconsistency has been documented previously. For example, in a dose-dependent study, half-dose almonds (37 ± 2 g/d) increased oleic acid in NEFA and TAG fractions; however, the full dose (75 ± 3 g/d) did not demonstrate an increase in NEFA fraction (57). Our dietary data indicates greater oleic acid intake in the almond group compared to the cracker group. The bioaccessibility of lipids from whole almonds during mastication and digestion may limit the amount of fat absorbed through the gastrointestinal tract (58), hence contributing to the incongruency between dietary intake and serum NEFA levels. However, since serum NEFA levels are predominately regulated by adipose triglyceride hydrolysis (59), these results suggest that almond consumption did not alter adipose composition or the rates of adipose lipolysis. Alternatively, because increased oleic acid can stimulate complete fatty acid oxidation (60), the increased intake with almond consumption may have increased oxidation including the oxidation of oleic acid itself to mask the potential to detect an increase in circulating levels. The greater levels of acetylcarnitine particularly at week 4 in the almond group suggests that beta-oxidation of fatty acids was increased and substantiates the data in the literature that increased oleic acid promotes beta oxidations of FFA. Nonetheless, our results suggest that in a free-living study, serum oleic acid may not be a viable biomarker of compliance with almond consumption.

Almond consumption also lowered the palmitoleic acid: palmitic acid ratio. A greater ratio is considered a diagnostic marker for early onset non-alcoholic steatohepatitis (61) and associated with the presence of impaired glucose tolerance and T2D (62) suggesting that the lower ratio with almond consumption may be indicative of a protective effect against metabolic disorders.

The targeted analysis reveals lower levels of omega-3 total fatty acids (TFAs) in the almond group. More specifically, almond consumption had lower levels of ALA, EPA (measured in NEFA and TFA fractions), ALA oxylipins (9-HOTE and 15(16)-EpODE), and DHEA, the acylethanolamide of the omega-3 derivative of DHA. Interestingly, our 24-hour dietary recall data revealed greater intake of EPA in the almond group compared to the cracker group and no differences in ALA intake by group suggesting that the estimated dietary intake of these fatty acids are not indicative of the actual changes in circulating levels. These discrepancies could be due to the low reproducibility of food omega-3 fatty acids by ASA24 (63). Furthermore, the complex metabolism of omega-3 fatty acids involves a series of enzymatic desaturation and elongation processes, interconversions between FAs, and translocation between different cellular compartments (64) that cannot be captured by estimating dietary intake.

The increases in aconitic, citric, isocitric, and succinic acids suggests that almond consumption stimulated the TCA cycle. The TCA cycle is a central metabolic pathway where key byproducts of nutrient digestion such as glucose, fatty acids, and some amino acids converge for energy production (65). The increased activity in the TCA cycle could be due to the higher dietary intake of MUFAs and PUFAs in the almond group. Higher fat intake has been found to elevate TCA cycle intermediates in rats (66) suggesting that increased availability of unsaturated FAs may increase the substrates for TCA cycle activity and in essence, feed the system. While in a recent human study, baseline levels of specific TCA cycle metabolites including isocitrate were associated with greater relative risk of T2D, the Mediterranean diet, a diet high in unsaturated fats, appeared to alleviate this risk (67). As mentioned previously, almond consumption increased acetylcarnitine, which is significant here because it is involved in the movement of acetyl-CoA into the mitochondria during beta-oxidation, thereby providing the raw materials for TCA cycle (68). Thus, the increase in TCA cycle activity may be facilitated by increased acetylcarnitine. These data provide a mechanism by which increased dietary MUFA (oleic acid) (and potentially PUFAs) can improve glucose metabolism (via enhanced TCA cycle activity). In the same subjects studied here, almond consumption improved glucose tolerance (21).

Another carnitine derivative, 3-hydroxyisovaleroylcarnitine, which is a degradation byproduct of ketogenic amino acids, was lower in the almond group. This may suggest that biotin intake was increased. Increased circulating levels of 3-hydroxyisovaleroylcarnitine is associated with impaired leucine catabolism due to reduced activity of 3-methylcrotonyl-CoA carboxylase, which is a biotin-dependent enzyme in asymptomatic, marginally biotin deficient adults (69, 70). Although ASA24 data does not report biotin values, nuts are considered good sources of biotin and the dietary results suggest that the consumption of biotin-rich foods (red meat and eggs) was increased in the almond group.

We have previously documented increased alpha-diversity of the gut microbiome with almond consumption (22). The present analysis shows an enrichment of amino acid biosynthesis and carbohydrate metabolism such as amino sugar and nucleotide sugar metabolism in the almond group at week 8. Our analyses revealed differential snack effects on the microbial community potential of several MIMOSA2-predicted amino acids such as glutamate, glutamine, tryptophan, phenylalanine, and proline (**Supplemental Table 2)**. However, the predicted microbial metabolism data are most useful when associated with the metabolomics data. The only compound that was positively predicted at the end of the intervention was N-acetyl-D-mannosamine (ManNAc), which is an acylaminosugar. The microbial community in the almond group at week 8 explained 14% of the variation in serum ManNAc. The analyses further suggest that sialic acid metabolism, which includes biosynthesis of N-acetylneuraminic acid (Neu5Ac) from ManNAc, and catabolism of Neu5Ac to ManNAc and subsequently to ManNAc-6-P, which ultimately enters glycolysis, are involved (71). Although serum ManNAc itself was not significantly different between the almond and cracker groups, we speculate that metabolomics and meta-transcriptomics of stool samples might reveal other interesting relationships associated with almond consumption.

Other potential products of microbial metabolism that were differentially influenced by diet included cyclo (Leu-Pro), indole-3-carboxaldehyde, phenylacetylglutamine, and pipecolic acid. Phenylacetylglutamine, which is a conjugate of glutamine and phenylacetate, the latter being derived from microbial metabolism of phenylalanine (72, 73), was greater in the almond group. Studies have documented the positive association of phenylacetylglutamine with alpha-diversity (72), which is supported by our study as well (data not shown). Pipecolic acid, which was lower in the almond group, could arise from food intake or produced by gut bacteria on lysine degradation (74). Although, cyclo (Leu-Pro) is a bacterium-derived dipeptide whose functional role isn’t well known, literature suggests that cyclodipeptides and their derivatives such as diketopiperazines, contribute to bacterial signaling systems (75–77). Almond consumption also resulted in greater indole-3-carboxaldehyde, which is a tryptophan metabolite that acts as a ligand for the aryl hydrocarbon receptor (AhR) in the intestinal immune cells that stimulate IL-22 production when activated. This activation is important for maintaining gut immunity (78), providing another potential benefit of almond consumption on gut health in addition to promoting alpha-diversity.

We used comprehensive analyses to study the serum metabolome of almond versus cracker consumers. The changes in microbial metabolism provide complementary insight into the metabolomics data. However, since we used prediction models for assessing microbial metabolism and community potential, the results should be interpreted with caution and in the context of the exploratory nature of the analyses. Moreover, more than half of the metabolites in our dataset could not be annotated owing to the nature of untargeted metabolomics, and biochemical relationships for most lipid mediators could not be defined due to the inherent limitations of KEGG database for lipidomic datasets. Nonetheless, these data provide significant insights on the effects of chronic almond consumption on metabolic pathways not previously explored. Future analyses will explore effects of factors such as BMI, sex, and metabolic risk on these outcomes.

Our results provide a deeper understanding of host TCA cycle and lipid metabolism with almond consumption in relatively healthy young adults. In addition, the findings also shed light into the interconnections between circulating metabolites and microbial metabolism in the context of an almond intervention. More generally, these findings provide further evidence for the potential impacts of dietary changes on host substrate metabolism and associated changes in gut microbe metabolism. Whether the changes in the gut microbe or metabolites influence host metabolism or vice versa remains to be elucidated but these data provide evidence for an association between gut microbe metabolism and host cellular metabolism.

## Data Availability

The datasets generated for this study are available on request to the corresponding author.

## AUTHOR RESPONSIBILITIES

JD and RMO designed and conducted the study. JWN and OF directed the metabolomics analyses and assisted with the data interpretation. JD drafted the manuscript. All authors revised the manuscript and approved the submitted version.

## REFERENCES

1. Rezzi S, Ramadan Z, Fay LB, Kochhar S. Nutritional Metabonomics: Applications and Perspectives. J Proteome Res American Chemical Society; 2007;6:513–25.

2. Grapov D, Adams SH, Pedersen TL, Garvey WT, Newman JW. Type 2 Diabetes Associated Changes in the Plasma Non-Esterified Fatty Acids, Oxylipins and Endocannabinoids. PLOS ONE 2012;7:e48852.

3. Adams SH, Hoppel CL, Lok KH, Zhao L, Wong SW, Minkler PE, Hwang DH, Newman JW, Garvey WT. Plasma acylcarnitine profiles suggest incomplete long-chain fatty acid beta-oxidation and altered tricarboxylic acid cycle activity in type 2 diabetic African-American women. J Nutr 2009;139:1073–81.

4. Fiehn O, Garvey WT, Newman JW, Lok KH, Hoppel CL, Adams SH. Plasma metabolomic profiles reflective of glucose homeostasis in non-diabetic and type 2 diabetic obese African-American women. PloS One 2010;5:e15234.

5. Gibbons H, O’Gorman A, Brennan L. Metabolomics as a tool in nutritional research. Curr Opin Lipidol 2015;26:30–4.

6. Cross AJ, Major JM, Sinha R. Urinary biomarkers of meat consumption. Cancer Epidemiol Biomark Prev Publ Am Assoc Cancer Res Cosponsored Am Soc Prev Oncol 2011;20:1107–11.

7. Rothwell JA, Fillâtre Y, Martin J-F, Lyan B, Pujos-Guillot E, Fezeu L, Hercberg S, Comte B, Galan P, Touvier M, et al. New Biomarkers of Coffee Consumption Identified by the Non-Targeted Metabolomic Profiling of Cohort Study Subjects. PLOS ONE Public Library of Science; 2014;9:e93474.

8. Heinzmann SS, Brown IJ, Chan Q, Bictash M, Dumas M-E, Kochhar S, Stamler J, Holmes E, Elliott P, Nicholson JK. Metabolic profiling strategy for discovery of nutritional biomarkers: proline betaine as a marker of citrus consumption. Am J Clin Nutr 2010;92:436–43.

9. Edmands WMB, Beckonert OP, Stella C, Campbell A, Lake BG, Lindon JC, Holmes E, Gooderham NJ. Identification of human urinary biomarkers of cruciferous vegetable consumption by metabonomic profiling. J Proteome Res 2011;10:4513–21.

10. Dhillon J, Ferreira CR, Sobreira TJP, Mattes RD. Multiple Reaction Monitoring Profiling to Assess Compliance with an Almond Consumption Intervention. Curr Dev Nutr 2017;1:e001545.

11. Andersen M-BS, Kristensen M, Manach C, Pujos-Guillot E, Poulsen SK, Larsen TM, Astrup A, Dragsted L. Discovery and validation of urinary exposure markers for different plant foods by untargeted metabolomics. Anal Bioanal Chem 2014;406:1829– 44.

12. Goodacre R. Metabolomics of a Superorganism. J Nutr Oxford Academic; 2007;137:259S–266S.

13. Marcobal A, Kashyap PC, Nelson TA, Aronov PA, Donia MS, Spormann A, Fischbach MA, Sonnenburg JL. A metabolomic view of how the human gut microbiota impacts the host metabolome using humanized and gnotobiotic mice. ISME J Nature Publishing Group; 2013;7:1933–43.

14. Kim Y, Keogh J, Clifton PM. Nuts and Cardio-Metabolic Disease: A Review of Meta-Analyses. Nutrients [Internet] 2018 [cited 2020 Jun 22];10. Available from: https://www.ncbi.nlm.nih.gov/pmc/articles/PMC6316378/

15. Garcia-Aloy M, Llorach R, Urpi-Sarda M, Tulipani S, Estruch R, Martínez-González MA, Corella D, Fitó M, Ros E, Salas-Salvadó J, et al. Novel Multimetabolite Prediction of Walnut Consumption by a Urinary Biomarker Model in a Free-Living Population: the PREDIMED Study. J Proteome Res American Chemical Society; 2014;13:3476–83.

16. Hernández-Alonso P, Cañueto D, Giardina S, Salas-Salvadó J, Cañellas N, Correig X, Bulló M. Effect of pistachio consumption on the modulation of urinary gut microbiota-related metabolites in prediabetic subjects. J Nutr Biochem 2017;45:48–53.

17. Tulipani S, Llorach R, Jáuregui O, López-Uriarte P, Garcia-Aloy M, Bullo M, Salas-Salvadó J, Andrés-Lacueva C. Metabolomics unveils urinary changes in subjects with metabolic syndrome following 12-week nut consumption. J Proteome Res 2011;10:5047–58.

18. Mora-Cubillos X, Tulipani S, Garcia-Aloy M, Bulló M, Tinahones FJ, Andres-Lacueva C. Plasma metabolomic biomarkers of mixed nuts exposure inversely correlate with severity of metabolic syndrome. Mol Nutr Food Res 2015;59:2480–90.

19. Dhillon J, Thorwald M, De La Cruz N, Vu E, Asghar SA, Kuse Q, Diaz Rios LK, Ortiz RM. Glucoregulatory and Cardiometabolic Profiles of Almond vs. Cracker Snacking for 8 Weeks in Young Adults: A Randomized Controlled Trial. Nutrients 2018;10:960.

20. Dhillon J, Li Z, Ortiz RM. Almond Snacking for 8 Weeks Increases Alpha-Diversity of the Gastrointestinal Microbiome and Decreases Bacteroides fragilis Abundance Compared to an Isocaloric Snack in College Freshmen. Curr Dev Nutr [Internet] [cited 2019 Jul 8]; Available from: https://academic.oup.com/cdn/advance-article/doi/10.1093/cdn/nzz079/5527932

21. Dhillon J, Thorwald M, De La Cruz N, Vu E, Asghar SA, Kuse Q, Diaz Rios LK, Ortiz RM. Glucoregulatory and Cardiometabolic Profiles of Almond vs. Cracker Snacking for 8 Weeks in Young Adults: A Randomized Controlled Trial. Nutrients 2018;10:960.

22. Dhillon J, Li Z, Ortiz RM. Almond Snacking for 8 wk Increases Alpha-Diversity of the Gastrointestinal Microbiome and Decreases Bacteroides fragilis Abundance Compared with an Isocaloric Snack in College Freshmen. Curr Dev Nutr [Internet] Oxford Academic; 2019 [cited 2020 Nov 16];3. Available from: https://academic.oup.com/cdn/article/3/8/nzz079/5527932

23. Dhillon J, Li Z, Ortiz RM. Almond Snacking for 8 wk Increases Alpha-Diversity of the Gastrointestinal Microbiome and Decreases Bacteroides fragilis Abundance Compared with an Isocaloric Snack in College Freshmen. Curr Dev Nutr [Internet] 2019 [cited 2019 Sep 17];3. Available from: https://academic.oup.com/cdn/article/3/8/nzz079/5527932

24. Fiehn O, Wohlgemuth G, Scholz M, Kind T, Lee DY, Lu Y, Moon S, Nikolau B. Quality control for plant metabolomics: reporting MSI-compliant studies. Plant J Cell Mol Biol 2008;53:691–704.

25. Kind T, Tolstikov V, Fiehn O, Weiss RH. A comprehensive urinary metabolomic approach for identifying kidney cancerr. Anal Biochem 2007;363:185–95.

26. Fiehn O. Extending the breadth of metabolite profiling by gas chromatography coupled to mass spectrometry. Trends Anal Chem TRAC 2008;27:261–9.

27. Fiehn O, Wohlgemuth G, Scholz M. Setup and Annotation of Metabolomic Experiments by Integrating Biological and Mass Spectrometric Metadata. In: Ludäscher B, Raschid L, editors. Data Integration in the Life Sciences. Berlin, Heidelberg: Springer; 2005. p. 224–39.

28. Scholz M, Fiehn O. SetupX--a public study design database for metabolomic projects. Pac Symp Biocomput 2007;169–80.

29. Matyash V, Liebisch G, Kurzchalia TV, Shevchenko A, Schwudke D. Lipid extraction by methyl-tert-butyl ether for high-throughput lipidomics. J Lipid Res 2008;49:1137– 46.

30. Tsugawa H, Cajka T, Kind T, Ma Y, Higgins B, Ikeda K, Kanazawa M, VanderGheynst J, Fiehn O, Arita M. MS-DIAL: data-independent MS/MS deconvolution for comprehensive metabolome analysis. Nat Methods Nature Publishing Group; 2015;12:523–6.

31. Sun Chongxiu, Alkhoury Kenan, Wang Ying I., Foster Greg A., Radecke Christopher E., Tam Kayan, Edwards Christina M., Facciotti Marc T., Armstrong Ehrin J., Knowlton Anne A., et al. IRF-1 and miRNA126 Modulate VCAM-1 Expression in Response to a High-Fat Meal. Circ Res American Heart Association; 2012;111:1054–64.

32. Smedes F. Determination of total lipid using non-chlorinated solvents. Analyst The Royal Society of Chemistry; 1999;124:1711–8.

33. Grapov D, Adams SH, Pedersen TL, Garvey WT, Newman JW. Type 2 Diabetes Associated Changes in the Plasma Non-Esterified Fatty Acids, Oxylipins and Endocannabinoids. PLOS ONE 2012;7:e48852.

34. Fan S, Kind T, Cajka T, Hazen SL, Tang WHW, Kaddurah-Daouk R, Irvin MR, Arnett DK, Barupal DK, Fiehn O. Systematic Error Removal Using Random Forest for Normalizing Large-Scale Untargeted Lipidomics Data. Anal Chem 2019;91:3590–6.

35. Barupal DK, Fiehn O. Chemical Similarity Enrichment Analysis (ChemRICH) as alternative to biochemical pathway mapping for metabolomic datasets. Sci Rep Nature Publishing Group; 2017;7:1–11.

36. Barupal DK, Haldiya PK, Wohlgemuth G, Kind T, Kothari SL, Pinkerton KE, Fiehn O. MetaMapp: mapping and visualizing metabolomic data by integrating information from biochemical pathways and chemical and mass spectral similarity. BMC Bioinformatics 2012;13:99.

37. Cao Y, Jiang T, Girke T. A maximum common substructure-based algorithm for searching and predicting drug-like compounds. Bioinformatics 2008;24:i366–74.

38. Shannon P, Markiel A, Ozier O, Baliga NS, Wang JT, Ramage D, Amin N, Schwikowski B, Ideker T. Cytoscape: a software environment for integrated models of biomolecular interaction networks. Genome Res 2003;13:2498–504.

39. Su G, Kuchinsky A, Morris JH, States DJ, Meng F. GLay: community structure analysis of biological networks. Bioinformatics Oxford Academic; 2010;26:3135–7.

40. Ye Y, Doak TG. A Parsimony Approach to Biological Pathway Reconstruction/Inference for Genomes and Metagenomes. PLOS Comput Biol Public Library of Science; 2009;5:e1000465.

41. Douglas GM, Maffei VJ, Zaneveld JR, Yurgel SN, Brown JR, Taylor CM, Huttenhower C, Langille MGI. PICRUSt2 for prediction of metagenome functions. Nat Biotechnol Nature Publishing Group; 2020;38:685–8.

42. Ritchie ME, Phipson B, Wu D, Hu Y, Law CW, Shi W, Smyth GK. limma powers differential expression analyses for RNA-sequencing and microarray studies. Nucleic Acids Res 2015;43:e47–e47.

43. Law CW, Chen Y, Shi W, Smyth GK. voom: precision weights unlock linear model analysis tools for RNA-seq read counts. Genome Biol 2014;15:R29.

44. Chong J, Liu P, Zhou G, Xia J. Using MicrobiomeAnalyst for comprehensive statistical, functional, and meta-analysis of microbiome data. Nat Protoc Nature Publishing Group; 2020;15:799–821.

45. Shaffer M, Thurimella K, Quinn K, Doenges K, Zhang X, Bokatzian S, Reisdorph N, Lozupone CA. AMON: annotation of metabolite origins via networks to integrate microbiome and metabolome data. BMC Bioinformatics 2019;20:614.

46. Dhillon J, Li Z, Ortiz RM. Almond Snacking for 8 Weeks Increases Alpha-Diversity of the Gastrointestinal Microbiome and Decreases Bacteroides fragilis Abundance Compared to an Isocaloric Snack in College Freshmen. Curr Dev Nutr [Internet] [cited 2019 Jul 27]; Available from: https://academic.oup.com/cdn/advance-article/doi/10.1093/cdn/nzz079/5527932

47. Borenstein Lab. MIMOSA2 [Internet]. 2020. Available from: http://borensteinlab.com/software_MIMOSA2.html

48. Noecker C, Eng A, Srinivasan S, Theriot CM, Young VB, Jansson JK, Fredricks DN, Borenstein E. Metabolic Model-Based Integration of Microbiome Taxonomic and Metabolomic Profiles Elucidates Mechanistic Links between Ecological and Metabolic Variation. mSystems [Internet] American Society for Microbiology Journals; 2016 [cited 2021 Mar 22];1. Available from: https://msystems.asm.org/content/1/1/e00013-15

49. Jambazian PR, Haddad E, Rajaram S, Tanzman J, Sabaté J. Almonds in the diet simultaneously improve plasma α-tocopherol concentrations and reduce plasma lipids. J Am Diet Assoc 2005;105:449–54.

50. Brigelius-Flohé R, Kelly FJ, Salonen JT, Neuzil J, Zingg J-M, Azzi A. The European perspective on vitamin E: current knowledge and future research. Am J Clin Nutr 2002;76:703–16.

51. Dimitrov NV, Meyer C, Gilliland D, Ruppenthal M, Chenoweth W, Malone W. Plasma tocopherol concentrations in response to supplemental vitamin E. Am J Clin Nutr 1991;53:723–9.

52. Reaven PD, Witztum JL. Comparison of supplementation of RRR-alpha-tocopherol and racemic alpha-tocopherol in humans. Effects on lipid levels and lipoprotein susceptibility to oxidation. Arterioscler Thromb J Vasc Biol 1993;13:601–8.

53. Law S-H, Chan M-L, Marathe GK, Parveen F, Chen C-H, Ke L-Y. An Updated Review of Lysophosphatidylcholine Metabolism in Human Diseases. Int J Mol Sci [Internet] 2019 [cited 2021 Mar 2];20. Available from: https://www.ncbi.nlm.nih.gov/pmc/articles/PMC6429061/

54. Heimerl S, Fischer M, Baessler A, Liebisch G, Sigruener A, Wallner S, Schmitz G. Alterations of Plasma Lysophosphatidylcholine Species in Obesity and Weight Loss. PLoS ONE [Internet] 2014 [cited 2021 Feb 25];9. Available from: https://www.ncbi.nlm.nih.gov/pmc/articles/PMC4207804/

55. Wallace M, Morris C, O’Grada CM, Ryan M, Dillon ET, Coleman E, Gibney ER, Gibney MJ, Roche HM, Brennan L. Relationship between the lipidome, inflammatory markers and insulin resistance. Mol Biosyst 2014;10:1586–95.

56. Zhang J-M, An J. Cytokines, Inflammation and Pain. Int Anesthesiol Clin 2007;45:27–37.

57. Nishi S, Kendall CWC, Gascoyne A-M, Bazinet RP, Bashyam B, Lapsley KG, Augustin LSA, Sievenpiper JL, Jenkins DJA. Effect of almond consumption on the serum fatty acid profile: a dose-response study. Br J Nutr 2014;112:1137–46.

58. Grundy MM, Lapsley K, Ellis PR. A review of the impact of processing on nutrient bioaccessibility and digestion of almonds. Int J Food Sci Technol 2016;51:1937–46.

59. Physiological regulation of NEFA availability: lipolysis pathway | Proceedings of the Nutrition Society | Cambridge Core [Internet]. [cited 2021 Apr 28]. Available from: https://www.cambridge.org/core/journals/proceedings-of-the-nutrition-society/article/physiological-regulation-of-nefa-availability-lipolysis-pathway/81DE2F9FCB84C6A38140D2D7EA63C5B7

60. Lim J-H, Gerhart-Hines Z, Dominy JE, Lee Y, Kim S, Tabata M, Xiang YK, Puigserver P. Oleic Acid Stimulates Complete Oxidation of Fatty Acids through Protein Kinase A-dependent Activation of SIRT1-PGC1α Complex. J Biol Chem 2013;288:7117–26.

61. Yamada K, Mizukoshi E, Seike T, Horii R, Terashima T, Iida N, Kitahara M, Sunagozaka H, Arai K, Yamashita T, et al. Serum C16:1n7/C16:0 ratio as a diagnostic marker for non-alcoholic steatohepatitis. J Gastroenterol Hepatol 2019;34:1829–35.

62. Lankinen MA, Stančáková A, Uusitupa M, Ågren J, Pihlajamäki J, Kuusisto J, Schwab U, Laakso M. Plasma fatty acids as predictors of glycaemia and type 2 diabetes. Diabetologia 2015;58:2533–44.

63. Yuan C, Spiegelman D, Rimm EB, Rosner BA, Stampfer MJ, Barnett JB, Chavarro JE, Subar AF, Sampson LK, Willett WC. Validity of a Dietary Questionnaire Assessed by Comparison With Multiple Weighed Dietary Records or 24-Hour Recalls. Am J Epidemiol 2017;185:570–84.

64. Dyall SC. Long-chain omega-3 fatty acids and the brain: a review of the independent and shared effects of EPA, DPA and DHA. Front Aging Neurosci [Internet] 2015 [cited 2021 Mar 10];7. Available from: https://www.ncbi.nlm.nih.gov/pmc/articles/PMC4404917/

65. Berg JM, Tymoczko JL, Stryer L. The Citric Acid Cycle. Biochem 5th Ed [Internet] W H Freeman; 2002 [cited 2021 Apr 1]; Available from: https://www.ncbi.nlm.nih.gov/books/NBK21163/

66. An Y, Xu W, Li H, Lei H, Zhang L, Hao F, Duan Y, Yan X, Zhao Y, Wu J, et al. High-Fat Diet Induces Dynamic Metabolic Alterations in Multiple Biological Matrices of Rats. J Proteome Res American Chemical Society; 2013;12:3755–68.

67. Guasch-Ferré M, Santos JL, Martínez-González MA, Clish CB, Razquin C, Wang D, Liang L, Li J, Dennis C, Corella D, et al. Glycolysis/gluconeogenesis- and tricarboxylic acid cycle–related metabolites, Mediterranean diet, and type 2 diabetes. Am J Clin Nutr 2020;111:835–44.

68. Randle PJ, Garland PB, Hales CN, Newsholme EA. The glucose fatty-acid cycle. Its role in insulin sensitivity and the metabolic disturbances of diabetes mellitus. Lancet Lond Engl 1963;1:785–9.

69. Stratton SL, Horvath TD, Bogusiewicz A, Matthews NI, Henrich CL, Spencer HJ, Moran JH, Mock DM. Plasma concentration of 3-hydroxyisovaleryl carnitine is an early and sensitive indicator of marginal biotin deficiency in humans1234. Am J Clin Nutr 2010;92:1399–405.

70. Horvath TD, Stratton SL, Bogusiewicz A, Pack L, Moran J, Mock DM. Quantitative Measurement of Plasma 3-Hydroxyisovaleryl Carnitine by LC-MS/MS as a Novel Biomarker of Biotin Status in Humans. Anal Chem American Chemical Society; 2010;82:4140–4.

71. Almagro-Moreno S, Boyd EF. Bacterial catabolism of nonulosonic (sialic) acid and fitness in the gut. Gut Microbes 2010;1:45–50.

72. Ottosson F, Brunkwall L, Smith E, Orho-Melander M, Nilsson PM, Fernandez C, Melander O. The gut microbiota-related metabolite phenylacetylglutamine associates with increased risk of incident coronary artery disease. J Hypertens 2020;38:2427–34.

73. Li M, Wang B, Zhang M, Rantalainen M, Wang S, Zhou H, Zhang Y, Shen J, Pang X, Zhang M, et al. Symbiotic gut microbes modulate human metabolic phenotypes. Proc Natl Acad Sci National Academy of Sciences; 2008;105:2117–22.

74. He M. Pipecolic acid in microbes: biosynthetic routes and enzymes. J Ind Microbiol Biotechnol 2006;33:401–7.

75. Ortiz-Castro R, Díaz-Pérez C, Martínez-Trujillo M, Río RE del, Campos-García J, López-Bucio J. Transkingdom signaling based on bacterial cyclodipeptides with auxin activity in plants. Proc Natl Acad Sci National Academy of Sciences; 2011;108:7253–8.

76. Holden MT, Ram Chhabra S, de Nys R, Stead P, Bainton NJ, Hill PJ, Manefield M, Kumar N, Labatte M, England D, et al. Quorum-sensing cross talk: isolation and chemical characterization of cyclic dipeptides from Pseudomonas aeruginosa and other gram-negative bacteria. Mol Microbiol 1999;33:1254–66.

77. Edlund A, Garg N, Mohimani H, Gurevich A, He X, Shi W, Dorrestein PC, McLean JS. Metabolic Fingerprints from the Human Oral Microbiome Reveal a Vast Knowledge Gap of Secreted Small Peptidic Molecules. mSystems [Internet] American Society for Microbiology Journals; 2017 [cited 2021 Apr 6];2. Available from: https://msystems.asm.org/content/2/4/e00058-17

78. Zhang LS, Davies SS. Microbial metabolism of dietary components to bioactive metabolites: opportunities for new therapeutic interventions. Genome Med [Internet] 2016 [cited 2021 Apr 7];8. Available from: https://www.ncbi.nlm.nih.gov/pmc/articles/PMC4840492/

