## Supplemental Files for "Almond Consumption for 8 Weeks Altered Host and Microbial Metabolism in Comparison to a Control Snack in Young Adults"

Supplemental Table 1: Enrichment analysis of differentially expressed (almond vs. cracker) microbial metabolic pathway genes at baseline and week 8 of the intervention.

| Baseline Almond vs. Cracker (39 mapped out of 59 differentially expressed genes) | | | | | |
| --- | --- | --- | --- | --- | --- |
| Pathway | Total | Expected | Hits | Pval* | FDR |
| Porphyrin and chlorophyll metabolism | 76 | 0.829 | 4 | 0.0087 | 0.648 |
| Propanoate metabolism | 82 | 0.895 | 4 | 0.0113 | 0.648 |
| Pyrimidine metabolism | 85 | 0.927 | 4 | 0.0128 | 0.648 |
| Ubiquinone and other terpenoid-quinone biosynthesis | 50 | 0.546 | 3 | 0.0164 | 0.648 |
| Biosynthesis of siderophore group nonribosomal peptides | 3 | 0.0327 | 1 | 0.0324 | 0.862 |
| Alanine, aspartate and glutamate metabolism | 65 | 0.709 | 3 | 0.0327 | 0.862 |
| Galactose metabolism | 71 | 0.775 | 3 | 0.041 | 0.925 |
| Starch and sucrose metabolism | 87 | 0.949 | 3 | 0.0676 | 1 |
| Synthesis and degradation of ketone bodies | 8 | 0.0873 | 1 | 0.0841 | 1 |
| Biosynthesis of amino acids | 223 | 2.43 | 5 | 0.0914 | 1 |
| Week 8 Almond vs. Cracker (113 mapped out of 142 differentially expressed genes) | | | | | |
| Pathway | Total | Expected | Hits | Pval* | FDR |
| Biosynthesis of amino acids | 223 | 6.8 | 17 | 0.000306 | 0.0279 |
| Amino sugar and nucleotide sugar metabolism | 126 | 3.84 | 12 | 0.000353 | 0.0279 |
| Starch and sucrose metabolism | 87 | 2.65 | 9 | 0.00112 | 0.0479 |
| Fructose and mannose metabolism | 88 | 2.68 | 9 | 0.00121 | 0.0479 |
| Lysine biosynthesis | 46 | 1.4 | 6 | 0.0024 | 0.0757 |
| Porphyrin and chlorophyll metabolism | 76 | 2.32 | 7 | 0.00769 | 0.203 |
| Pyruvate metabolism | 86 | 2.62 | 7 | 0.0148 | 0.299 |
| Cyanoamino acid metabolism | 32 | 0.976 | 4 | 0.0152 | 0.299 |
| Cysteine and methionine metabolism | 78 | 2.38 | 6 | 0.0301 | 0.482 |
| Biosynthesis of ansamycins | 1 | 0.0305 | 1 | 0.0305 | 0.482 |

*Hypergeometric tests conducted in MicrobiomeAnalyst.

Supplemental Table 2: Metabolites showing significant differences in the MIMOSA2 predicted microbial community metabolic potential (CMP) scores

| Metabolite | emmean Almond | SE Almond | emmean Cracker | SE Cracker | P-value* |
| --- | --- | --- | --- | --- | --- |
| L-Glutamate | -0.62 | 0.13 | 0.22 | 0.13 | 2.70E-05 |
| L-Glutamine | 2.03 | 0.12 | 0.95 | 0.12 | 1.60E-08 |
| L-Tryptophan | 1.20 | 0.09 | -0.27 | 0.10 | 7.35E-17 |
| L-Phenylalanine | 1.38 | 0.06 | -0.51 | 0.07 | 2.47E-31 |
| Fumarate | -1.60 | 0.14 | -2.06 | 0.15 | 2.81E-02 |
| Myo-Inositol | -0.33 | 0.08 | 1.22 | 0.09 | 2.42E-20 |
| L-Proline | 1.32 | 0.08 | -0.49 | 0.08 | 5.77E-26 |
| 5'-Methylthioadenosine | -1.10 | 0.09 | 0.24 | 0.10 | 1.07E-14 |
| Maltose | 0.60 | 0.12 | -0.17 | 0.12 | 2.21E-05 |
| L-Kynurenine | -0.07 | 0.13 | 0.61 | 0.14 | 8.62E-04 |
| D-Sorbitol | -0.32 | 0.12 | 0.11 | 0.13 | 1.72E-02 |
| Hydroxyproline | -1.36 | 0.08 | 0.54 | 0.09 | 3.55E-25 |
| Myo-Inositol 4-phosphate | 0.33 | 0.08 | -1.22 | 0.09 | 2.42E-20 |

*Baseline-adjusted linear mixed model analysis on CMP scores of metabolites

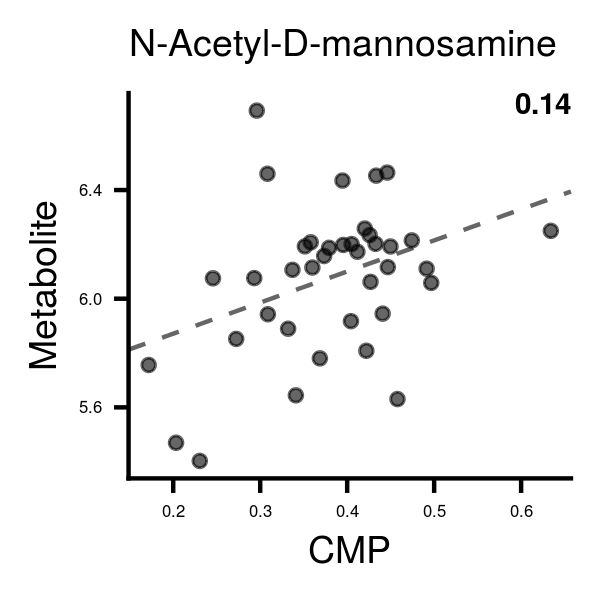

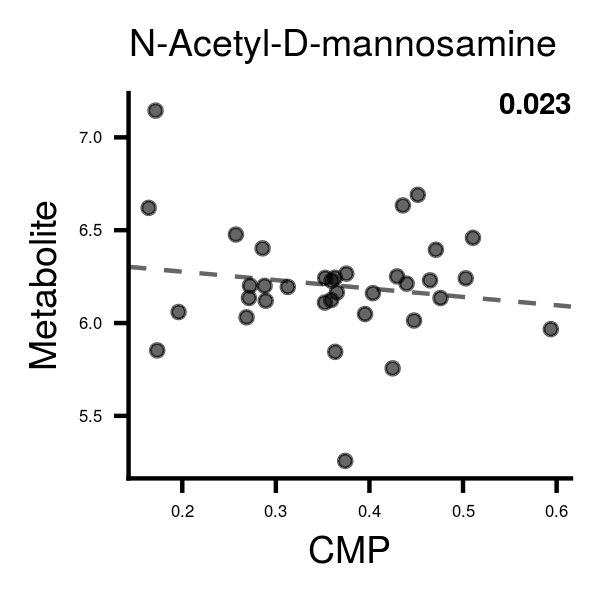

**Almond	Cracker**

R^2^= 0.14

R^2^= 0.023

Supplemental Figure 1: Association of serum N-acetyl-D-mannosamine with its MIMOSA2 predicted community metabolic potential (CMP) score.

Y-axis is log-transformed serum metabolite levels. P<0.05 for difference between correlation coefficients.

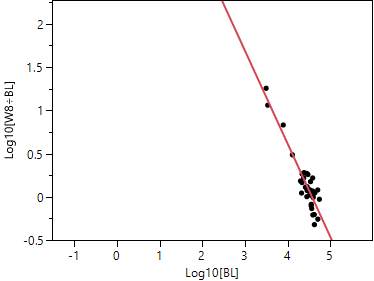

Almond group

R^2^= 0.86

Supplemental Figure 2: Changes in alpha-tocopherol expressed as a function of baseline concentrations. The magnitudes of concentration changes with almond consumption decrease as baseline concentrations increase indicating the presence of a saturable pool. BL, baseline; W8, week 8. ÷
